# IMPACT OF SEXUALITY IN PATIENTS WITH MOTOR NEURON DISEASE

**DOI:** 10.1101/2025.06.10.25329362

**Authors:** R. Yolanda Morgado Linares, Íñigo Rojas-Marcos, Marco Mesa, Eva Martínez, Macarena Cabrera-Serrano, Carmen Paradas

**Author notes:** Correspondance autor: Rosalía Yolanda Morgado Linares, Hospital Virgen Macarena, Doctor Fedriani 3. 41009, Sevilla (Spain), Carmen Paradas, Hospital Virgen del Rocio. Calle Manuel Siurot sn. 41013, Sevilla (Spain).

## Abstract

**Background:** Sexuality is a relevant yet understudied aspect in patients with motor neuron diseases (MND). This study aimed to assess whether sexuality in MND patients is influenced by motor, cognitive, behavioral, or mood disturbances, and to explore whether being sexually active impacts caregiver burden.

**Methods:** We conducted a cross-sectional observational study involving 70 patients with MND and their primary caregivers from three ALS multidisciplinary units in Seville, Spain. Data were collected between October 2018 and July 2021. Functional, cognitive, behavioral, and mood assessments were conducted using ALSFRSr, ECAS, and HADS scales, respectively. Sexuality was evaluated through gender-specific questionnaires. Caregiver burden was assessed using the Zarit Burden Interview. All of these scales have their validated Spanish version.

**Results:** Among 66 patients who completed the sexuality assessment, 42.9% reported sexual activity in the previous month. Of these, 70% had no sexual dysfunction, while 26.7% had moderate and 3.3% severe dysfunction. No correlation was found between sexual activity and motor function (ALSFRSr) or caregiver burden. Sexual activity was less frequent in patients with apathy (p=0.026), depression (p=0.021), or frontotemporal dementia (p=0.013). Among sexually active patients, those with cognitive impairment—particularly memory dysfunction—had higher rates of arousal (p=0.021) and orgasm difficulties (p=0.019). No other clinical parameters were related to sexuality. Sexuality did not influence caregiver burden either.

**Conclusions:** Sexuality in MND is more associated with cognitive and mood factors than with motor disability. Addressing sexuality in clinical practice may contribute to improving quality of life, and should be integrated into the multidisciplinary care of MND patients.

## INTRODUCTION

Motor neuron diseases (MNDs) are a group of neurodegenerative diseases of uncertain aetiology in most cases, being hereditary in 5-10% of cases. Characterized by degeneration of the pyramidal pathway, they classically include: primary lateral sclerosis (PLS) (exclusive involvement of the first motor neuron), progressive muscular atrophy (PMA) (exclusive involvement of the second motor neuron), progressive bulbar palsy (PBP) and amyotrophic lateral sclerosis (ALS) (involvement of first and second motor neuron). Although it is now accepted that this is a continuous spectrum with Frontotemporal Dementia(1) MNDs, with ALS as their main representative, are considered as “rare diseases”, with low incidence and prevalence (conditioned by rapid evolution in most cases), whose epidemiological study is not easy. In general terms, an incidence of 1.6-1.7/100,000 inhabitants per year(2) and a prevalence of 4.42/100,000 inhabitants are estimated(2,3).

Sexuality in people with MND is an aspect that has long been ignored. In the few studies available, 90%(4) or 100%(5) of patients report that they have never been asked about sexuality, probably due to the devastating nature of the disease that has focused attention on motor aspects. In a disease for which there is still no curative treatment, day-to-day management aims to improve quality of life. Sexuality is one of the areas that can influence quality of life.

Despite of the scarcity of bibliography about the topic as reflected in a 2019 review(6), it seems that sexuality is an issue of interest to patients with ALS and their caregivers(7,8), and it usually influences the quality of life(9–11) for both patients and caregivers.

In general, studies in this area consist of single descriptions of severe dysfunction in advanced cases (12), compilations of unstructured interviews(9,13) or the application of non-validated scales or surveys specifically designed for the study(7,12,14), as well as comments from patients or caregivers(8,11). However, we have identified only one study(15) that used validated questionnaires for both men and women to assess the sexuality of neurological patients, including nine ALS patients. The published bibliography identifies physical factors such as fatigue, pain or weakness (7,9,11) as interfering with sexuality. It also suggests that the sexuality of MND patients does not differ significantly from that of their healthy counterparts in the same age group(7). Additonally, it notes that that sexuality problems in MND patients are similar to those described for other neuromuscular diseases in adulthood(15). No bibliography has been found regarding the relationship between sexuality and cognitive impairment in patients with MND. To our knowledge, no large-scale studies with an appropriate methodology to assess sexuality in MND have been performed.

We decided to further investigate sexuality in MND as one of the conditioning areas of quality of life. The study aims to assess whether sexuality is influenced by patients’ motor, cognitive, behavioural or mood disturbances, and whether the fact that the patient is sexually active influences the burden on the primary caregiver.

## PATIENTS AND METHODS

### Patients

We conducted a cross-sectional observational study of a series of cases. The study received approval from the Local Ethic Committee of the Hospital Universitario Virgen Macarena/Virgen del Rocío (Seville). Patients eligible for the study were selected from the ALS Units of the three main hospitals in Sevilla, namely Hospital Universitario de Valme, Hospital Universitario Virgen del Rocío, Hospital Universitario Virgen Macarena. Patient inclusion commenced on October 1st, 2018 and concluded on July 31st, 2021. The inclusion period was extended due to the exceptional health situation caused by the SarsCov2 pandemic, thus, no patients were enrolled in the study from March 2020 to May 2021. None of the patients in the study had SarsCov2 infection.

In most cases, patients signed informed consent themselves. For patients who were unable to sign due to mobility issues, verbal consent was obtained either directly or through a communication device, and it was signed by their primary caregiver.

### Inclusion criteria

1. - Patients over the age of 18 who have been evaluated in one of the three ALS Multidisciplinary Units in Seville and who meet the revised El Escorial(16). diagnostic criteria for possible, probable, or definite ALS.
2. - Patients over 18 years of age, treated in one of the three ALS Multidisciplinary Units in Seville diagnosed with other variants of MND in any of its forms of presentation: PLS, PMA or PBP.
3. - Patients whose clinical situation allowed the study examinations to be carried out either through oral communication or the use of alternative communication devices (laser pointer, eye reader or other writing devices).

### Exclusion criteria

1. - Patients who do not sign or revoke the informed consent for the study.
2. - Patients affected by neoplasia or other concomitant disease that may influence or affect the course of the disease.
3. - Patients who are in a clinical trial with pharmacological intervention. 4.- Patients lacking an effective communication system.

### Variables of the study

The study collected demographical data, clinical functionality measured by the revised Amyotrophic Lateral Sclerosis Functional Rating Scale(17) (ALSFRSr), cognitive and behavioral state measured by the Edinburgh Cognitive and Behavioral ALS Screen (ECAS)(18), mood disorders measured by the Hospital Anxiety and Depression Scale (HADS)(19) and caregiver burden measured by the Zarit Burden Interview(20) to the main caregiver. All of these scales have their validated Spanish versions.

### Sexuality questionnaires

Two versions of the questionnaire have been used to assess sexuality.

The “Questionnaire on the sexual function of women”, validated in Spanish(21), was used for women. To ensure the assessment of comparable areas, an ad hoc questionnaire was created for male subjects based on the female version.

Although the questionnaires are designed to be self-administered, due to the characteristics of the disease that make it difficult in most cases to fill out the documents, the questionnaire was administered by the professional who conducted the visit. Caregiver were not present during the visit to allow the patient the necessary privacy to complete the questionnaire and thus to ensure reliability in the responses.

The first question in the sexuality questionnaire is a key question:

> *“Have you been sexually active in the last 4 weeks? (Informative note. Sexual activity can be with a partner or through one’s own sexual stimulation, including caresses, games, penetration, masturbation, etc*.*)”*; If the answer to this question is negative, the questionnaire will not continue.

The rest of the survey assesses six domains that are considered evaluative of sexual activity, and other domains that add descriptive information but do not influence the diagnosis of sexual dysfunction.

The diagnostic domains for sexual dysfunction include: desire, arousal, lubrication (for women only), orgasm, difficulties with vaginal penetration (for women only), anticipatory anxiety, overall sexual satisfaction, sexual initiative, degree of sexual communication and erectile dysfunction (for men only).

The descriptive domains are: characteristics of sexual activity, frequency of sexual activity and existence or not of a sexual partner.

Following the normative data of the questionnaire, scores above 50% in every domain assessed was considered within normality. Scores below 50% in at least one of the evaluative domains were considered indicative of sexual dysfunction. Punctuations between 50%-26% were considered as moderate dysfunction and puntuations below 25% indicate severe dysfunction.

### Statistics

This article is part of a larger study that examined multiple variables in patients with motor neuron disease and their relationships with each other. The sample size was calculated based on the main objective of investigating the relationship between cognitive impairment and functional status. A sample size of n=26 was required. The calculation was also performed with some of the secondary objectives, such as cognitive impairment and caregiver burden, for which a sample size of n=45 was estimated.

Quantitative variables were described using means and standard deviations if the distribution was symmetrical, or medians and quartiles in the case of asymmetrical distributions. Qualitative variables were expressed as frequencies and percentages. The normality of the distributions was verified using the Shapiro-Wilk test. The comparison of numerical variables between two groups was performed using the Student’s t-test or the non-parametric Mann-Whitney U test. Numerical variables between two groups were compared using the Student’s t-test or the nonparametric Mann-Whitney U test. Significant differences were quantified with 95% confidence intervals. Quantitative variables were compared using one-way ANOVA or Kruskal-Wallis (depending on whether or not they followed a normal distribution). Associations between qualitative variables were studied by creating contingency tables and applying the Chi-square test or non-asymptotic methods. Correlations between numerical parameters were analyzed by calculating Spearman’s non-linear correlation coefficients. In the event of evidence of high-degree associations, logistic regression models were generated. For all tests performed, bilateral statistical significance was set at p≤0.05. Missing values for the variables studied were omitted from the analysis. Data analysis was performed using IBM SPSS 28 statistical software.

We used th STROBE reporting checklist(22) when editing this manuscript, included in supplement A

## RESULTS

Seventy patients and their 70 main caregivers were included in the study. Clinical and demographic data are described in tables 1 and 2.

**Table 1.** Clinical and demographic data

|  |  |  |
| --- | --- | --- |
| Sex |  | 61.4% Men<br>38.6% Women |
| Age |  | Mean 61.10 CI (58.51 – 63.69) |
| Familiar ALS |  | 10% |
| Sporadic ALS |  | 90% |
| Phenotype |  | 80% ALS<br>10% PLS<br>10% PMA |
| ALS-FTD |  | 7 patients (10%) |
| ALSFRS<sub>r</sub> |  | Me 35 (IQR 25-40) |
| Communication | Verbal | 87.1% |
|  | Alternative | 12.9% |
| Movement | Walk | 61.4% |
|  | Wheelchair | 38.6% |
| Non invasive ventilation |  | 28.6% |
| Tracheostomy |  | 0% |
| Gastrostomy |  | 10% |

Me: Median, IQR: Interquartile range (P25-P75)

**Table 2.**
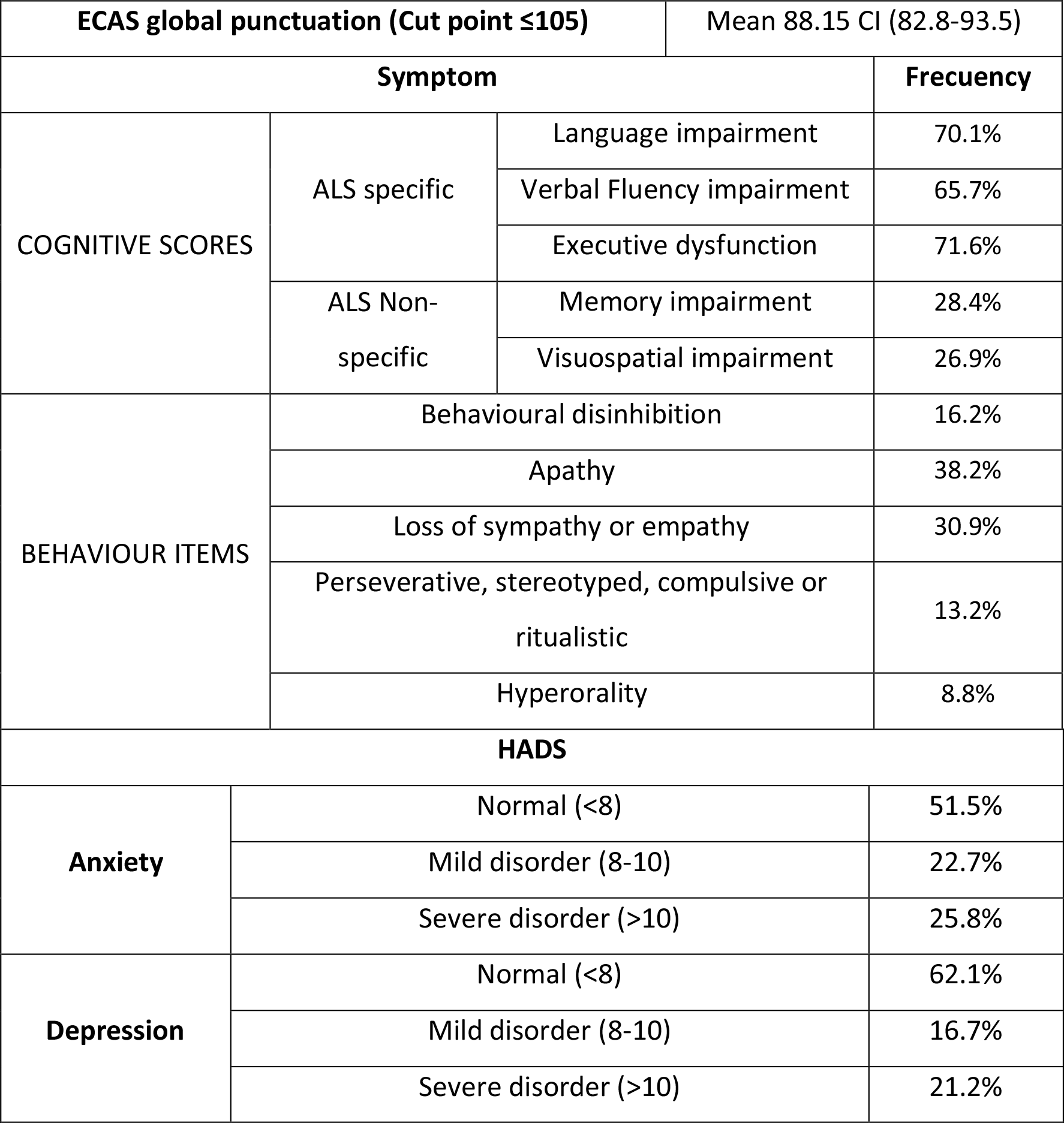
Results of the ECAS and HADS scales

| ECAS global punctuation (Cut point ≤105) |  |  | Mean 88.15 CI (82.8-93.5) |
| --- | --- | --- | --- |
| Symptom |  |  | Frecuency |
| COGNITIVE SCORES | ALS specific | Language impairment | 70.1% |
|  |  | Verbal Fluency impairment | 65.7% |
|  |  | Executive dysfunction | 71.6% |
|  | ALS Non-specific | Memory impairment | 28.4% |
|  |  | Visuospatial impairment | 26.9% |
| BEHAVIOUR ITEMS | Behavioural disinhibition |  | 16.2% |
|  | Apathy |  | 38.2% |
|  | Loss of sympathy or empathy |  | 30.9% |
|  | Perseverative, stereotyped, compulsive or ritualistic |  | 13.2% |
|  | Hyperorality |  | 8.8% |
| HADS |  |  |  |
| Anxiety | Normal (<8) |  | 51.5% |
|  | Mild disorder (8-10) |  | 22.7% |
|  | Severe disorder (>10) |  | 25.8% |
| Depression | Normal (<8) |  | 62.1% |
|  | Mild disorder (8-10) |  | 16.7% |
|  | Severe disorder (>10) |  | 21.2% |

### Sexuality (Questionnaire on the sexual function of women & version adapted for men)

Four patients were unable to complete the sexuality questionnaire due to their clinical situation, which forced them to conclude the visit prematurely. The results of the sexuality questionnaire are shown for the 66 patients that completed the assessment. Thirty-six patients (23 males and 13 females) did not have sexual activity in the previous month to the visit and the questionnaire was finalized after the first key question. Thirty patients (42.9%) answered that they had sexual activity in the last month, so they completed questionnaire (11 women and 19 men).

Eight patients (26,7%) had moderate sexual dysfunction and 1 (3,3%) severe sexual dysfunction, the rest (70%) did not present sexual dysfunction. Ninety percent of patients were satisfied with their sexual activity and 86.7% were satisfied with their sexuality. Other descriptive measures of the sample are summarized in table 3.

**Table 3.** Results of the Sexual Function Questionnaire

| **Patients who had been sexually active in the previous 4 weeks and completed the questionnaire (n= 30)** |  | **Frequency** |
| --- | --- | --- |
| **Presence of a partner** |  | **93.3%** |
| **Communication with the partner** | Without problem | 81.5% |
|  | Moderate communication | 3.7% |
|  | Without communication | 14.8% |
| **Frequency of sexual activity** | 1-2 times a month | 50% |
|  | 3-4 times a month | 26.7% |
|  | 5-8 times a month | 10% |
|  | 9-12 times a month | 3.3% |
|  | >12 times a month | 10% |
| **Initiative in the sexual relationship** | Never | 28.6% |
|  | Sometimes | 17.9% |
|  | Often | 21.4% |
|  | Almost always- always | 32.1% |
| **Anticipatory anxiety** | Yes | 3.3% |
|  | No | 96.7% |
| **Orgasm** | No alterations | 83.3% |
|  | Moderate disorder | 6.7% |
|  | Severe disorder | 10% |
| **Desire** | No alterations | 80% |
|  | Moderate disorder | 13.3% |
|  | Severe disorder | 6.7% |
| **Excitement** | No alterations | 96.7% |
|  | Severe disorder | 3.3% |
| **Pain** | No alterations | 96,7% |
|  | Severe disorder | 3,3% |

Next, we analysed sexual activity of MND patients and the results of the questionnaire in those sexually active patients, in relation to other aspects of the disease and the burden of caregivers:

### Cognitive and behavioural status (ECAS)

#### Cognitive impairment

No differences were found in ECAS score or in any of the cognitive aspects, including memory impairment, language impairment, verbal fluency impairment, executive dysfunction, visuospatial impairment, between individuals who had intercourse in the previous four weeks and those who had not.

#### Behaviuor

None of the patients that fulfilled frontotemporal dementia criteria(23) had sexual activity in the previous four weeks (p=0.013). Additionally, patients with apathy were found to be less sexually active (p=0.026). No other behavior alteration made any difference in sexual activity.

In the sexually active group that completed the questionnaire, we found that patients with lower global punctuation on the ECAS assessment, presented more frequently arousal impairment (p<0.001) and troubles in the communications with their partners (p=0.016).

More specifically, patients with memory impairment also had more frequent problems in the arousal phase (p=0.021) and orgasm (p=0.019). No other cognitive or behavioural aspects determined any difference in the phases of sexual activity.

### Functional situation (ALSFRSr)

There was no significant correlation found between sexuality and the ALSFRSr, either in the presence or absence of sexual activity, or in the results of the questionnaire in sexually active patients.

The same absence of relationship between sexuality and functional status was found when the functional status was measured by items as the communication (verbal or alternative) and the need of wheelchair, mechanical ventilation or gastrostomy.

### Mood disturbances (HADS)

Patients with higher score in the depression subscale were less sexually active (p=0.021). No other differences were found in relation to mood alterations and sexuality.

### Caregiving burden (ZARIT)

No statistically significant differences were observed in the burden of caregivers depending on whether or not the patient had been sexually active in the previous month.

## DISCUSSION

Sexuality can have a significant impact on a patient’s quality of life. This prompted us to conduct the present study. However, there is a lack of attention given to this area of study, as evidenced by the limited number of studies available in the literature(6). Furthermore, the previous studies have applied a wide range of different methods to assess sexuality, making them incomparable and precluding general conclusions.

In this study, 94.3% of patients participated in the sexuality questionnaire, with only four patients unable to complete the interview due to their advanced functional clinical situation, not because they refused to discuss the topic of sexuality. The high participation rate indicates a strong willingness to discuss the topic of sexuality among patients. In the same line, other studies show that sexuality is an important issue for the majority of patients with MND, even in advanced stages of the disease(11,13). However, studies such as the one conducted by Wasner et al.(7) in the German population, have a response rate of only 68%. The main reasons given by patients or caregivers for not participating were “issue unimportant” or “issue inconvenient”. To compare with the general population, we have examined at the most recent survey on sexuality in the general population conducted by the National Institute of Statistics in Spain in 2003(24). The survey targeted individuals between the ages of 18 and 49, and 10,980 interviews were conducted. The rate of refusal to respond was 30.3%, which is similar to that of the German study in ALS patients.

Education and cultural aspects can influence when dealing with sexuality, which is traditionally a topic that neither doctors nor patients systematically include in the medical interview. A 2017 study conducted in the New York health area(14) found that almost half of the participating doctors reported feeling uncomfortable discussing sexuality with their patients. Cultural aspects are undoubtedly significant in the treatment of sexuality even from a medical point of view. However, it is essential to consider other factors as well. The high participation rate in our study may be attributed to the fact that the interview was part of a larger study that assessed various scales, including sexuality. The participants’ trust in their reference neurologist may have also played a role. All these aspects should be taken into account when designing the interviews in future studies that address sexuality in patients with MND.

The present analysis revealed that the majority of patients in our sample exhibited no symptoms of sexual dysfunction defined as alterations in any of the phases of sexual activity, including desire, arousal, orgasm, or the presence of pain, with 70% displaying no alterations, while only 3.3% exhibited severe dysfunction and 26.7% mild dysfunction. These results contrast sharply with those reported Nasimbera et al(15) who evaluated sexuality in different neurological diseases in an Argentine population, including nine patients with ALS. In that study severe sexual dysfunction was observed in five patients (55.6%), moderate in two (22.2%), and no dysfunction in the remaining two. The authors hypothesise that these disorders may be associated with motor disability, ventilatory disturbances and depressive symptoms. However, the high prevalence of sexual dysfunction in the aforementioned study may be influenced by the relatively small sample size, which limits the generalizability of the findings. In contrast, our study, with a sample more than three times larger, provides a more robust estimate of the frequency of sexual dysfunction in this population.

To the best of our knowledge, there has been no prior research on the satisfaction of MND patients with their sexuality. In our study, we found that 90% of the sexually active patients, reported being satisfied with their sexual activity. In the distribution by sex, in our sample, 100% of men and 72.7% of women feel satisfied. These percentages are well above the 40% of women or 60% of men who were satisfied with their sexuality in the Nasimbera study(15), which included several neurological diseases, besides nine ALS patients, or the 45% reported in Germany among people with chronic disease or disability(25). The level of sexual satisfaction reported in our study is comparable to that of the healthy Argentine population(15), and exceeds that described in the general German population(25) and in the population over 50 years of age in San Diego(26) (USA). Therefore, our study suggest that the disease does not have a negative impact on the perception of satisfaction with sexuality. It is likely that geographic and cultural factors probably influence the perception of sexuality. For this reason, our sample is more similar to the data obtained from Latino populations.

In relation to the factors that can influence sexuality, we have not found a relationship with the motor functional situation measured with the ALSFRSr. Previous studies analyzing sexuality in relation to functional situation of MND patients are scarce. Atkins et al (27) analysed the couple relationship (not limited to sexual relationships) in ALS patients after the diagnosis. They performed functional assessment using the “ALS Severity Scale”, and concluded that psychosocial factors have a greater influence on the couple relationship than factors related to the symptoms of the disease. Other studies(8,9) are merely descriptive compilations of interviews stating that intimate relationships are affected by physical issues such as fatigue, pain, interference with devices or weakness. In Wasner’ study(7), despite the fact that they describe the patients’ ALSFRSr, the correlation of the alterations in the sphere of sexuality was not carried out with the said scale, but with the subjective assessment of the participants, and they mentioned the weakness as one of the main problems in sexual activity, although they did not apply any type of statistical methodology. In contrast, our study supports that motor symptoms or physical limitations do not significantly interfere with patients’ sexuality.

Our study found that cognitive factors can interfere with sexuality. There were no differences in the ECAS score, either overall score or when broken down into cognitive areas, between sexually active and inactive individuals. However, when considering only sexually active individuals, a relationship between cognitive impairment and certain aspects of sexual activity, such as arousal and communication with a partner, is observed. These aspects are impaired in patients with cognitive impairment. Memory impairment is the main factor associated with sexual dysfunction. Patients with memory impairment experience more arousal and orgasm problems. There was no association found between the parameters assessed in the sexuality questionnaires and language impairment, verbal fluency, executive dysfunction, or visuospatial impairment. As we have not found bibliography about the relationship between sexuality and cognitive impairment in patients with MND, we have looked at studies that assess sexuality in other neurodegenerative diseases that occur with dementia(28,29) or in the elderly population(26) and they reflect the influence of cognitive impairment on sexuality, which is in line with our study.

Among behavior disorders, it was observed that patients with apathy were less sexually active than those without apathy. Similarly, patients who fulfilled FTD criteria also showed reduced sexual activity. There are references to sexuality in patients with ALS-FTD that emphasize the appearance of inappropriate sexual behavior and disinhibition(30). Our work includes disinhibition in the ECAS in a general way (not only in relation to sexual behavior) and we did not obtain a relationship between the presence or absence of disinhibition and alteration in sexuality, as with the rest of the behavioural disorders studied.

Considering mood disorders, depression - but not anxiety-has been observed to lead to a reduction in sexual activity. This has also been observed in the general elderly population(26), where individuals with depression (even paucisymptomatic) experience a decrease in the frequency of sexual activity, satisfaction, and communication with their partners, as well as in other neurodegenerative diseases(15) where a correlation between depression and sexual dysfunction has been identified.

In conclusion, our study uncovers very interesting data in relation to the sexuality of MND patients, which has not been previously analyzed in depth. We have identified that the alterations in sexuality are more related to cognitive aspects than to motor alterations. Additionally, the frequency of sexual activity decreases when apathy or depressive symptoms appear. Our work is pioneering in this regard, and is also, to our knowledge, the first to focus on the sexuality of patients with MND in Spain.

Sexuality has been shown to be an influential element in quality of life, which, in the absence of curative treatment, is the main objective in people with MND. Therefore, addressing sexuality is important both in the recognition of a potential problem and in the establishment of measures of improvement, and should be more actively treated in the multidisciplinary teams. Targeting cognitive and emotional factors may contribute to enhanced sexual well-being and, by extension, overall quality of life. Continued research is warranted to elucidate these relationships and inform the development of tailored interventions.

## Data Availability

All data produced in the present study are available upon reasonable request to the authors

## DATA AVAILABILITY

The data that support the findings of this study are available from the corresponding author, RYML, upon reasonable request.

## DECLARATION OF FOUNDING

No funding was received.

## AUTHORS CONTRIBUTION

CP and RYML designed the study. RYML, IR-M, MM, EM, MC-S and CP provided and cared for study patients. RYML and MM made the clinical studies to the patients, interviewed the main caregivers and collected data. MC-S provided critical discussion on the research. CP and RYML supervised and mentored all work. RYML coordinated all the study and wrote the initial manuscript. All authors contributed to the final version of the manuscript.

## ACKNOWLEDGEMENTS

The authors would like to express their gratitude to the patient associations ‘ELA Andalucía’ and ‘Saca la lengua a la ELA’ for their valuable collaboration, as well as to all the patients and caregivers who participated in the study.

The authors would like to thank Carmen, Nuria and Pilar, nurse case managers at the hospitals where the study was conducted, the patient associations “ELA Andalucía” and “Saca la lengua a la ELA” for their valuable collaboration, as well as all the patients and caregivers who participated in the study.

